# COVID-19 patient serum less potently inhibits ACE2-RBD binding for various SARS-CoV-2 RBD mutants

**DOI:** 10.1101/2021.08.20.21262328

**Authors:** Daniel Junker, Alex Dulovic, Matthias Becker, Teresa R. Wagner, Philipp D. Kaiser, Bjoern Traenkle, Katharina Kienzle, Stefanie Bunk, Carlotta Struemper, Helene Haeberle, Kristina Schmauder, Natalia Ruetalo, Nisar Malek, Karina Althaus, Michael Koeppen, Ulrich Rothbauer, Juliane S. Walz, Michael Schindler, Michael Bitzer, Siri Göpel, Nicole Schneiderhan-Marra

**Author notes:** denotes shared senior authorship and corresponding authors. Contact Information Nicole Schneiderhan-Marra –. Email Address Postal Address – Markwiesenstrasse 55, 72770 Reutlingen, Germany. Siri Göpel –. Email Address Postal Address – Otfried-Müller-Strasse 10, 72076 Tübingen, Germany.

## Abstract

As global vaccination campaigns against SARS-CoV-2 proceed, there is particular interest in the longevity of immune protection, especially with regard to increasingly infectious virus variants. Neutralizing antibodies (Nabs) targeting the receptor binding domain (RBD) of SARS-CoV-2 are promising correlates of protective immunity and have been successfully used for prevention and therapy. As SARS-CoV-2 variants of concern (VOCs) are known to affect binding to the ACE2 receptor and by extension neutralizing activity, we developed a bead-based multiplex ACE2-RBD inhibition assay (RBDCoV-ACE2) as a highly scalable, time-, cost-, and material-saving alternative to infectious live-virus neutralization tests. By mimicking the interaction between ACE2 and the RBD, this serological multiplex assay allows the simultaneous analysis of ACE2 binding inhibition to the RBDs of all SARS-CoV-2 VOCs and variants of interest (VOIs) in a single well. Following validation against a classical virus neutralization test and comparison of performance against a commercially available assay, we analyzed 266 serum samples from 168 COVID-19 patients of varying severity. ACE2 binding inhibition was reduced for ten out of eleven variants examined compared to wild-type, especially for those displaying the E484K mutation such as VOCs beta and gamma. ACE2 binding inhibition, while highly individualistic, positively correlated with IgG levels. ACE2 binding inhibition also correlated with disease severity up to WHO grade 7, after which it reduced.

## Introduction

Neutralizing antibodies (Nabs) prevent infection of the cell with pathogens or foreign particles by neutralizing them, eliminating a potential threat and rendering the pathogen or particle harmless [1]. The longevity of a Nab response has important implications for immune protection and vaccination strategies. In SARS-CoV-2, Nabs interfere with the cell entry mechanism primarily by blocking the interaction of the receptor binding domain (RBD) with the human cell receptor angiotensin converting enzyme 2 (ACE2) [2,3]. The RBD of SARS-CoV-2 is target of approximately 90 % of the neutralizing activity present in immune sera [4], with a lack of Nabs correlating with risk of fatal outcome [5,6]. Passive transfer of Nabs through convalescent serum or as monoclonal antibodies have been shown to provide protection from infection [7-9], with several Nabs drugs granted emergency use authorization by the U.S. Food and Drug Administration [10-13].

Since the first documented infections in Wuhan China [14], SARS-CoV-2 has continually evolved, with the emergence of global variants of concern (VOCs) being of particular importance. As of this moment, the WHO lists the alpha (B.1.1.7) [15], beta (B.1.351) [16], gamma (P.1) [17], delta (B.1.617.2) [18] and omicron (B.1.1.529) [19] strains as VOCs [20], in addition to further variants of interest (VOIs) such as lambda (C.37)[21]. The emergence and disappearance of variants and continual mutation of SARS-CoV-2 is of particular relevance for vaccine development, as all currently licensed vaccines [22-25] only elicit an immune response against the spike protein based on the original Wuhan-Hu-1 isolate (hereon referred to as “wild-type”) [26,27]. Several studies have already found that both convalescent and post-vaccinated sera have lower neutralization capacities against beta and gamma VOCs [28-30]. Of particular concern are mutations on amino acid residue (aa) 484 (e.g. E484K), which seem to confer escape from vaccine control, with an additional mutation on aa 501 (e.g. N501Y) increasing this effect [31].

In order to lead development of new vaccines and safely lift social restrictions, definitive correlates of protective immunity are necessary [32]. The gold standard for Nabs assessment are virus neutralization tests (VNTs), however these require live infectious virions which must be handled in biosafety level 3 (BSL3) laboratories, as well as access to variant strains of SARS-CoV-2. In this study we developed and applied RBDCoV-ACE2, a multiplex ACE2-RBD inhibition assay based upon the antibody-mediated inhibition of ACE2-RBD binding. This automatable assay enables simultaneous screening of serum samples for the presence of Nabs against a great number of VOCs/VOIs in a single well, making it a time-, material- and cost-effective alternative to live VNTs or classical ELISAs. Following in-depth validation of the assay, we analyzed the IgG antibody response and ACE2 binding inhibition of 266 serum samples from 168 COVID-19 patients with mild to severe disease progression towards eleven different SARS-CoV-2 variant RBDs including the alpha, beta, gamma and delta VOCs.

## Results

### ACE2-RBD inhibition assay (RBDCoV-ACE2) validation

To investigate the inhibition of ACE2 binding by SARS-CoV-2 VOCs, we developed and established a high-throughput bead-based multiplex ACE2-RBD inhibition assay (from here on referred to as “RBDCoV-ACE2”). This assay mimics the ACE2-RBD interaction and thereby detects the presence of Nabs against SARS-CoV-2 that inhibit this interaction. At the time of experimentation, RBDCoV-ACE2 contained the RBDs of SARS-CoV-2 wild-type and 11 different variants (alpha, beta, gamma, epsilon, eta, theta, kappa, delta, lambda, Cluster 5 and A.23.1).

To validate the assay, we both compared performance to a standard VNT (**Figure 1**), as well as completed technical validation to FDA bioanalytical guidelines verifying reagent stability, assay precision, freeze-thaw stability and parallelism (**Figure S1**). An assay validation sample set of 16 samples (12 convalescent, 4 pre-pandemic) was measured by VNT against wild-type and with RBDCoV-ACE2. The results of both assays showed a strong correlation (Spearman’s rank 0.95), confirming that RBDCoV-ACE2 is measuring neutralizing antibodies specifically (**Figure 1**). Technical validation performed with a set of 6 samples (4 vaccinated, 1 infected, 1 pre-pandemic) confirmed that RBDCoV-ACE2 is highly reproducible, as seen by the low intra- and inter-assay variation (all CVs under 5% and 7% respectively, **Figure S1a and b, Table S3**). ACE2 buffer was shown to be stable both in storage (4°C) and at room temperature (21 °C), with minimal loss in performance compared to freshly prepared buffer (**Figure S1c**). Similarly, the biotinylated ACE2 stock solution showed high freeze-thaw stability (all CVs under 13%, **Figure S1d**). Parallelism was used to optimize the assay conditions and to ensure that the ACE2 concentration was in linear range (**Figure S1e**). Percentage coefficients of variation (%CV) of all technical validation experiments for every analyzed samples are summarized in **Table S3**. Lastly, to ensure that the multiplex nature of the assay was not causing competition between beads for ACE2 which would have resulted in artificially deflated values, the assay was performed as both a singleplex (for all VOCs) and multiplex with 24 samples (19 COVID-19 infected, 5 pre-pandemic), with no difference in performance between the two bead compositions found (**Figure S2**).

**Figure 1.**
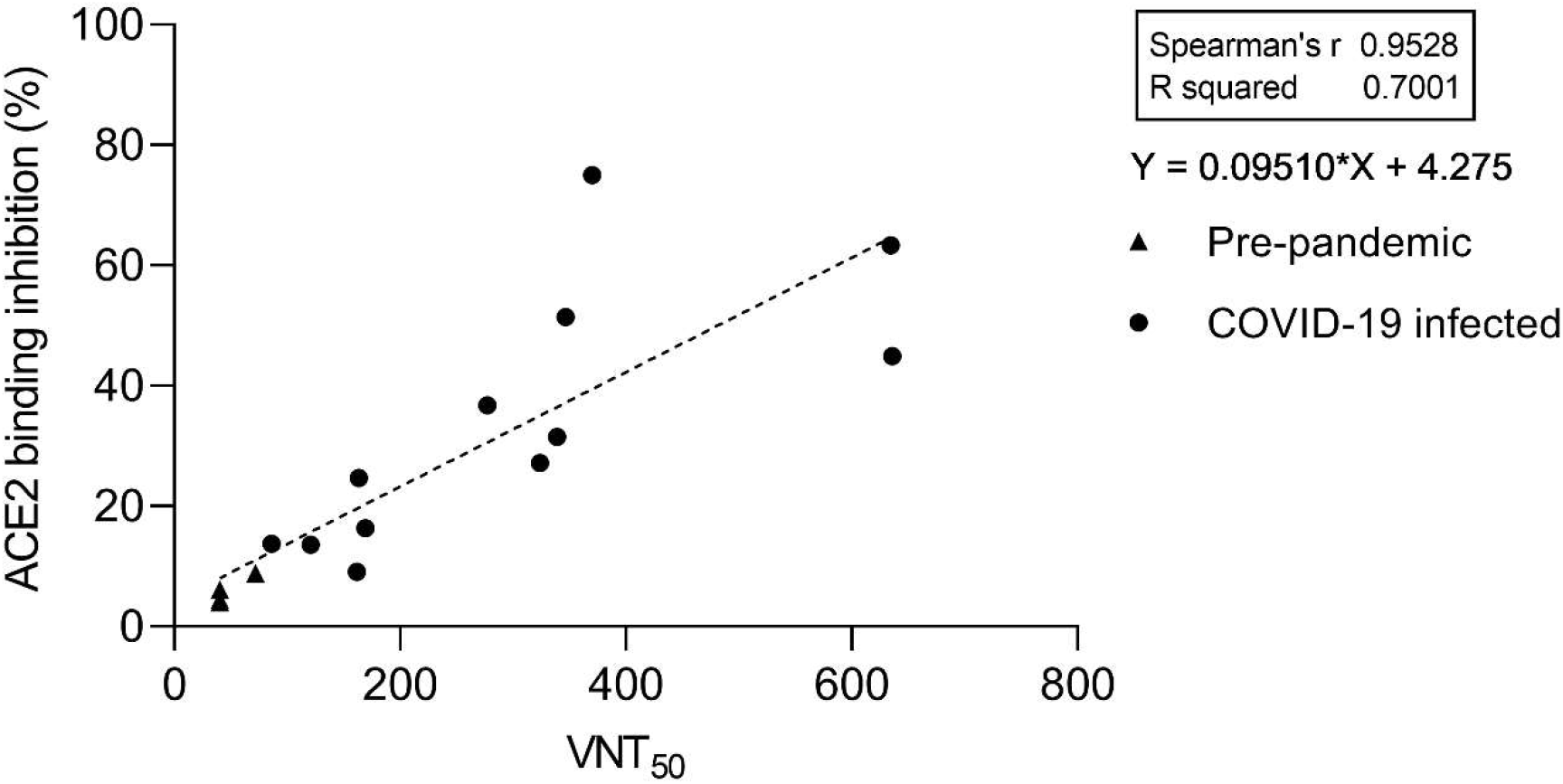
Comparison between RBDCoV-ACE2 and a virus neutralization test (VNT). Serum samples (n=16) of pre-pandemic (n=4) and COVID-19 convalescent (n=12) individuals were measured using both assays and analyzed by linear regression. The equation of the dashed regression line is shown next to the graph. VNT results are depicted as half-maximal inhibiting serum dilutions (VNT_50_), RBDCoV-ACE2 results are shown in percentage inhibition of ACE2 binding. Correlation analysis was performed after Spearman and the correlation coefficient r is shown.

### RBDCoV-ACE2 comparison to commercially available assay

To compare RBDCoV-ACE2 performance to a similar commercially available inhibition assay, we initially tested our assay validation sample set on NeutraLISA and compared its performance to the VNT (**Figure 2a**). While the results of the two assays did correlate (Spearman’s rank 0.94), the NeutraLISA appeared to reach a plateau and saturate, as seen by the high inhibition percentage for all samples with a VNT50 greater than 350. To confirm this plateau effect, we analyzed a subset of samples from our COVID-19 sample collection on both RBDCoV-ACE2 and the NeutraLISA, finding that while a strong correlation between the results existed (Spearman’s rank 0.84, **Figure 2b**), the saturation plateau was still present (**Figure 2c**). This suggests that RBDCoV-ACE2 has a more dynamic range and better resolution, especially in the higher inhibition percentages. When classifying samples as being either positive or negative, samples with a inhibition percentage under 20 % are considered negative for the NeutraLISA [33]. As both assays detect bound ACE2, we implemented a similar cut-off for RBDCoV-ACE2. Overall, 30.4% of samples (51/168) were considered negative in both assays, while a further 55.4% (93/168) were considered positive in both (**Figure 2b**). Of the remaining samples, 4 (2.4 %) exceeded 20 % binding inhibition only in RBDCoV-ACE2, while 20 (11.9 %) exceeded 20 % inhibition in the NeutraLISA only. Overall the stronger correlation between RBDCoV-ACE2 and VNT (**Figure 1**) compared to NeutraLISA and VNT, as well as the increased dynamic range, proves RBDCoV-ACE2 has superior assay performance.

**Figure 2.**
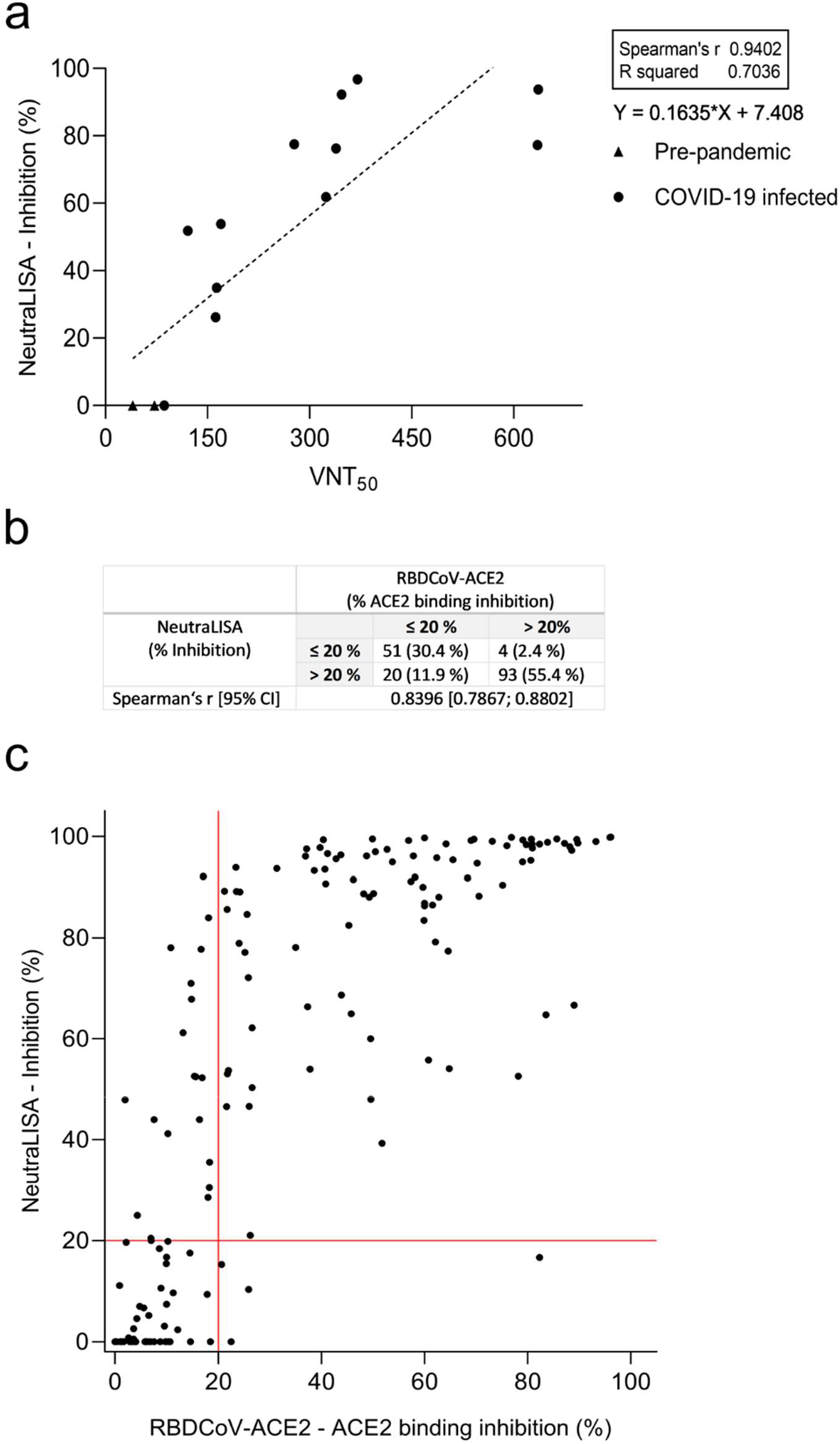
Correlation between SARS-CoV-2 NeutraLISA and VNT and comparison to RBDCoV-ACE2. (a) Correlation and linear regression between NeutraLISA and VNT results for pre-pandemic (n=4) and COVID-19 infected (n=12) samples. Correlation analyses were performed after Spearman and correlation coefficients r are shown. (b) Descriptive statistics of the (c) correlation between NeutraLISA and RBDCoV-ACE2. One sample from each individual (n=168) was measured using both assays correlation was calculated after Spearman. Samples were classified as being negative (non-neutralizing) if they had a value below 20% (red lines).

### ACE2 binding inhibition is reduced for mutant RBDs

Having developed and validated RBDCoV-ACE2, as well as identifying superior performance to a commercially available kit, we then analyzed ACE2 binding inhibition within 266 serum samples from 168 COVID-19 patients (COVID-19 sample collection), including longitudinal samples from 35 donors. Samples were measured against RBD wild-type and 11 variants (hereafter referred to as “RBD mutants”) of SARS-CoV-2. All RBD mutants except A.23.1 showed decreased ACE2 binding inhibition compared to wild-type (1.2-fold (Cluster 5) to 14.1-fold (beta), **Figure 3**) in serum samples taken within the first 49 days post initial positive PCR test. In the set of tested VOCs, alpha had the lowest reduction in ACE2 binding inhibition (1.2-fold), followed by delta (1.5-fold), gamma (6.4-fold) and beta (14.1-fold). While reduction in ACE2 binding inhibition was variant-specific, mutations at critical residues (e.g. E484K) appeared to have the largest effect (**Figure 3**). Among the current and former VOIs, epsilon had the lowest decrease (1.4 fold), followed by lambda (2.3 fold), kappa (3.3 fold), eta (5.7 fold) and theta (9.0 fold).

**Figure 3.**
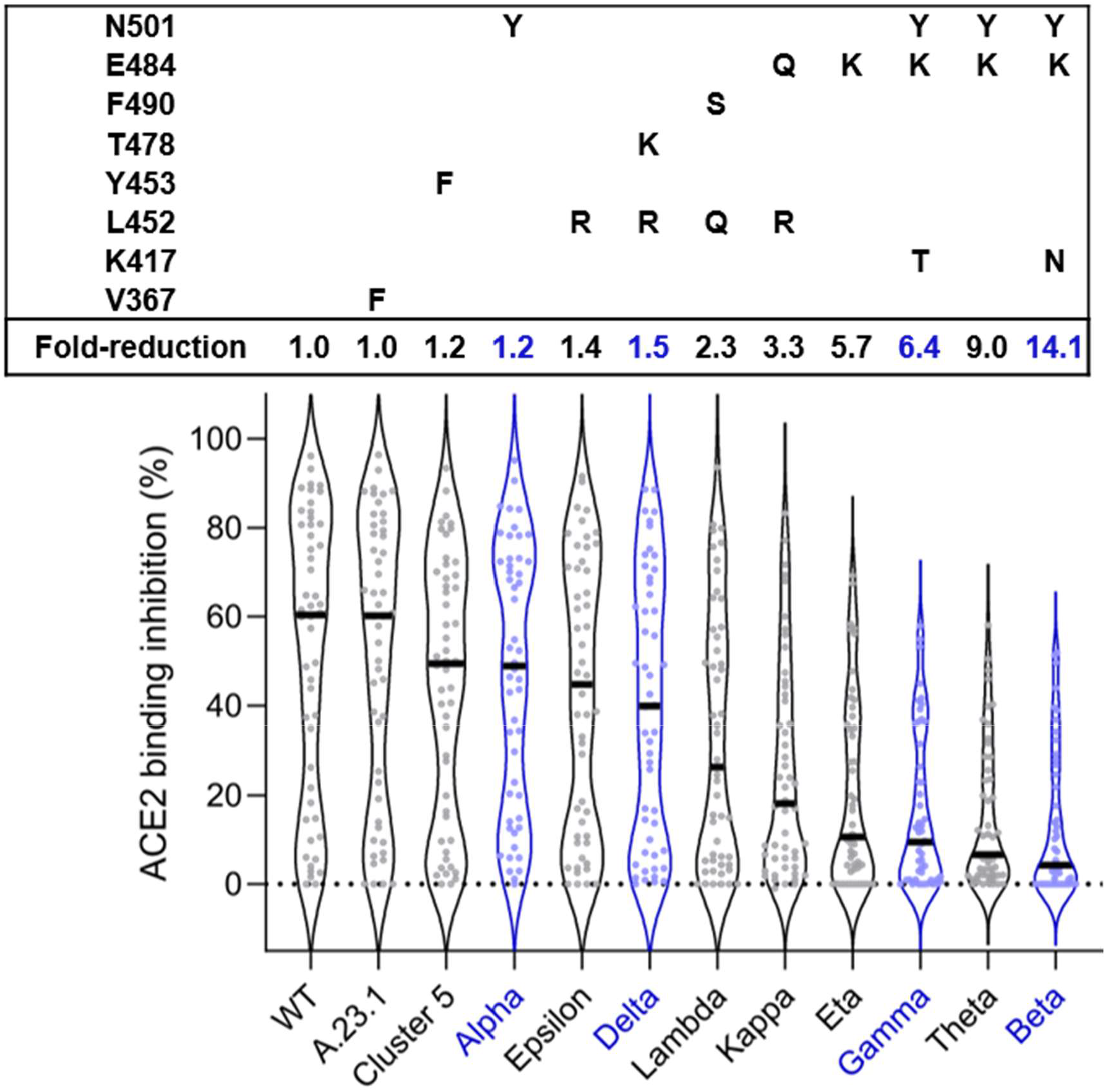
ACE2 binding inhibition varies between RBD mutants. Violin plots showing ACE2 binding inhibition (%) of individual serum samples from 7 to 49 days post PCR (n=50, depicted as dots) against RBD mutants. Black horizontal lines represent medians. Fold-decrease of ACE2 binding inhibition in comparison to wild-type corresponds to the ratio between the medians of wild-type and the respective RBD mutant. VOC-RBDs are shown in blue. Mutations of each RBD mutant are shown in the box above the violin plot.

### ACE2 binding inhibition correlates with antibody production against spike domains

To determine if a correlation existed between ACE2 binding inhibition and RBD-specific antibody levels, we analyzed all samples with MULTICOV-AB [34]. ACE2 binding inhibition and SARS-CoV-2 RBD IgG antibody responses were positively correlated (all Spearman’s correlation coefficients above 0.70, **Figure 4**) with variant-specific differences still present and reflecting. Additionally, we could show the positive correlation between ACE2 binding inhibition and S1 / trimeric spike antibody production (**Figure S3a and b**). The ACE2 binding inhibitions of both S1 and trimeric spike coated beads compared to the inhibition of RBD beads were strongly correlated (all Spearman’s correlation coefficients above 0.95) (**Figure S3c and d**). In contrast, beads coated with the SARS-CoV-2 spike S2 domain which does not interact with ACE2 *in vivo* showed no ACE2 binding in our assay. Those findings confirm specific binding of ACE2 to its natural binding partners and therefore reaffirms that the presence of neutralizing antibodies is being detected. For all RBD mutants, the increase in ACE2 binding inhibition most commonly occurred once IgG RBD MFI levels exceeded 10,000 (**Figure 4**). Notably, there was individual variation among the samples, with some having high ACE2 binding inhibition but relatively low IgG responses. For RBD mutants with a E484K mutation (eta, gamma, theta and beta), more than 78 % of all samples were considered negative, compared to 42 % for wild-type (**Figure 4**).

**Figure 4.**
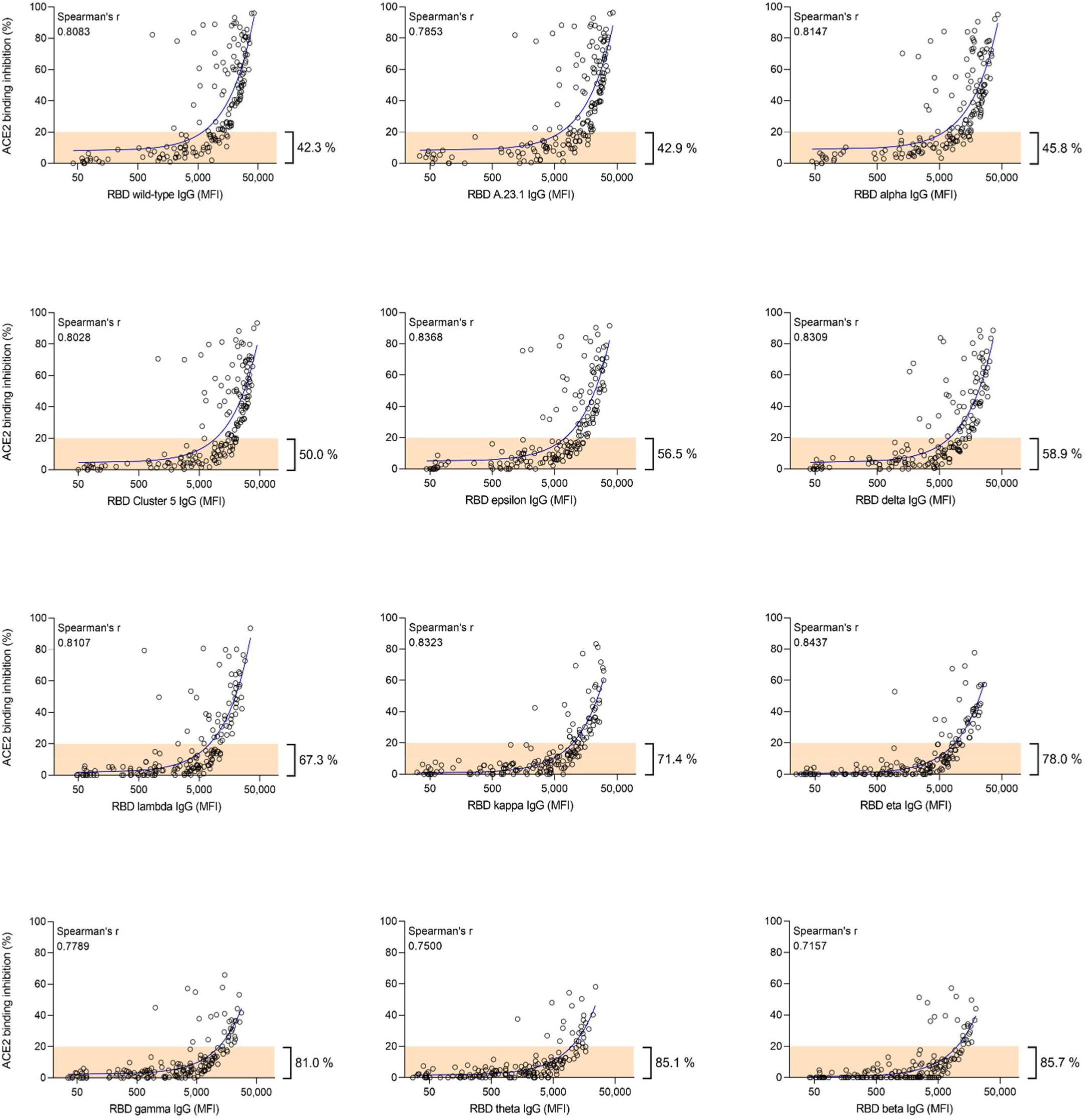
Correlation between anti-RBD IgG MFI signals and ACE2 binding inhibition (%) of serum samples from COVID-19 patients for wild-type and 11 RBD mutants. Regression analysis comparing ACE2 binding inhibition (%) and IgG responses (MFI) for wild-type and all RBD mutants included in the study. Each circle represents one sample (n=168). For longitudinal donors with more than one sample available, the sample closest to 20 days post positive PCR diagnosis was selected. The percentage next to the bracket indicates the proportion of samples with ACE2 binding inhibition ≤ 20% (in orange). Spearman’s correlation coefficient (r) is specified for every correlation.

### ACE2 binding inhibition decreases over time

To examine whether ACE2 binding inhibition changes over time, we analyzed longitudinal samples from 35 study participants (range 1-290 days post-initial positive PCR). ACE2 binding inhibition and RBD antibody titers originally remained low directly following a positive test, before rapidly increasing (mean peak at day 23 post-PCR) and then decreasing **(Figure 5a, b**). Due to the highly individualistic nature of the responses, we confirmed this pattern by analyzing a subset of six individuals with similar sample collection points (**Figure 5c, d**). As delta represents the current dominant global strain, we then examined whether any differences in ACE2 binding inhibition and antibody binding were present within this variant compared to wild-type. Overall, ACE2 binding inhibition and IgG response followed the same pattern for all samples as for wild-type (**Figure 5e, f**). We then confirmed that this pattern was true for all RBD variants (**Figure 5g, h**). As expected, while there were differences in reduction in binding inhibition between the variants, all variants examined follow the same pattern of reduced binding inhibition over time.

**Figure 5.**
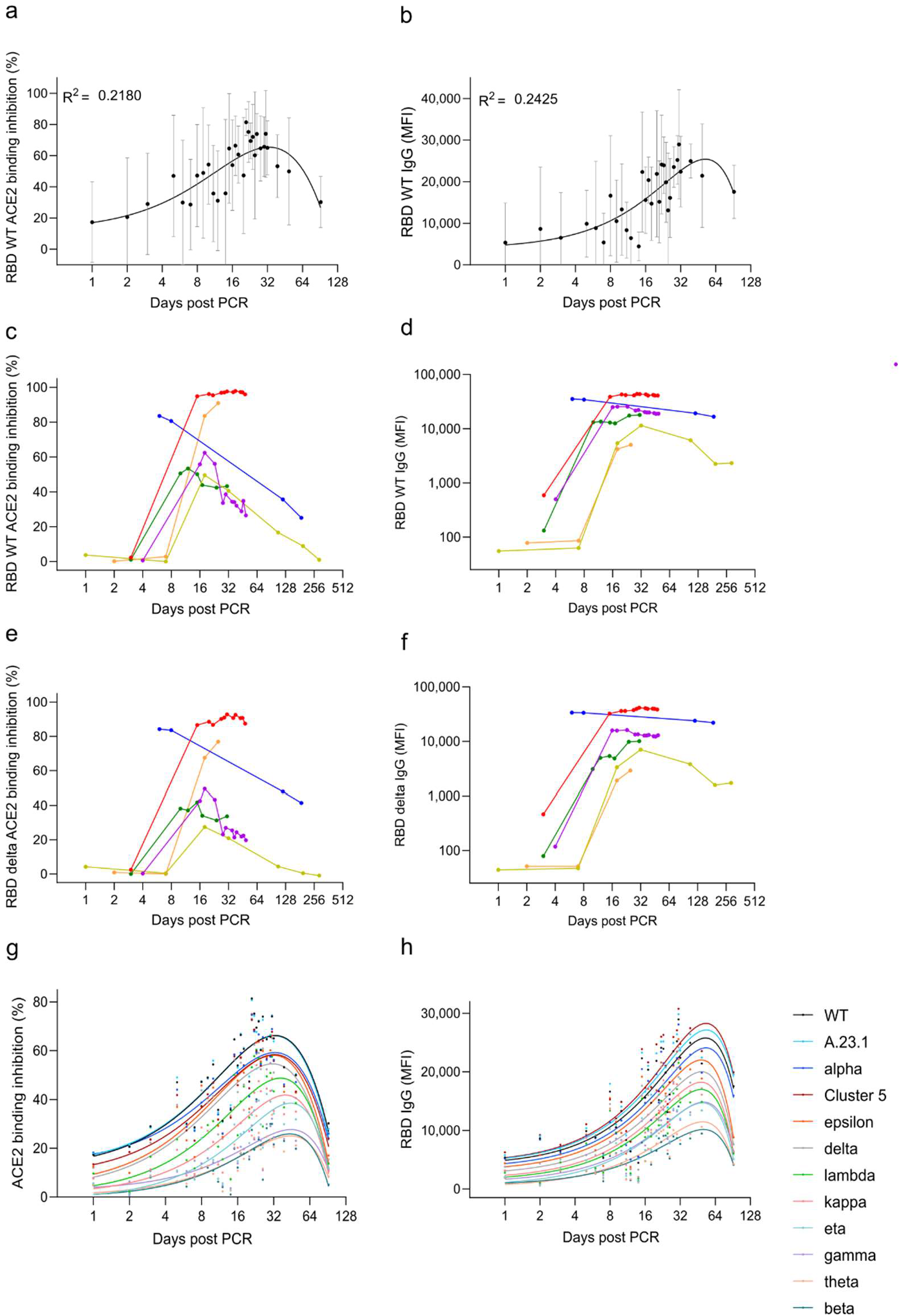
Longitudinal analysis of ACE2 binding inhibition and anti-RBD IgG levels in Covid-19 patients. Mean ACE2 binding inhibition (%) and IgG responses (MFI) for wild-type RBD against time post positive PCR test for samples (n=149) taken from 1 to 92 days post PCR are shown (a, b). Black dots indicate mean responses with standard deviation indicated by the error bars. The same analysis is then shown for longitudinal samples of selected donors (n=6) for wild-type (c, d) and RBD delta (e, f). For all RBD mutants, mean ACE2 binding inhibition (%) and mean IgG responses (MFI) 1 to 92 days post PCR included in the study is shown (g, h). Each variant is illustrated by a different color according to the figure key.

### ACE2 binding inhibition correlates with disease severity

We then examined correlations between ACE2 binding inhibition and COVID-19 disease severity within our population of COVID-19 patients. The severity of COVID-19 infection was determined according to the WHO grading scale (see methods). For analysis purposes, samples were split into two separate timeframes, 7-49 days post-initial positive PCR and ≥ 50 days post-initial positive PCR, in order to examine both the log and lag stages of infection. While all WHO grades (except for 5 and 8 for samples ≥ 50 days post-initial positive PCR) were represented within both timeframes, the early log timeframe consisted mostly of samples in WHO grades 4 and 6, while the later lag timeframe samples were mostly WHO grades 2 to 4. ACE2 binding inhibition was examined for both WT and delta to confirm no differences between wild-type and the variants existed. Regardless of timeframe, ACE2 binding inhibition increased as disease severity increased. Within the early log timeframe, ACE2 binding inhibition for wild-type and RBD delta increased steadily with disease severity up to grade 7 (WHO grading scale, hospitalized patients needing intubation and mechanical ventilation), before decreasing for patients of grade 8 (fatal disease course) (**Figure 6a, b**). Within the later lag timeframe, ACE2 binding inhibitions increased with disease severity (**Figure 6c, d**), however there was an overall reduction for grades 4 to 7 compared to the early timeframe for both wild-type and delta (**Figure 6c, d**). As expected, anti-RBD IgG levels also correlated with disease severity in both timeframes for wild-type and delta (**Figure 6e-h)**. Peak mean IgG levels were observed at grade 6 severity for wild-type and grade 7 for delta 7 – 49 days post PCR. Post 49 days, mean IgG levels peaked for patients with grade 6 severity. As confirmation, confounding variables (age, gender, BMI) were examined for any potential effect on the results (**Figure S4**). While gender had no effect, we did find correlations between ACE2 binding inhibition and donor age for samples taken ≥ 50 days post-positive PCR (p = 0.0001), as well as BMI for samples collected in both timeframes (< 49 days p = 0.0330, ≥ 50 days p = 0.0017) (**Figure S4d, f**).

**Figure 6.**
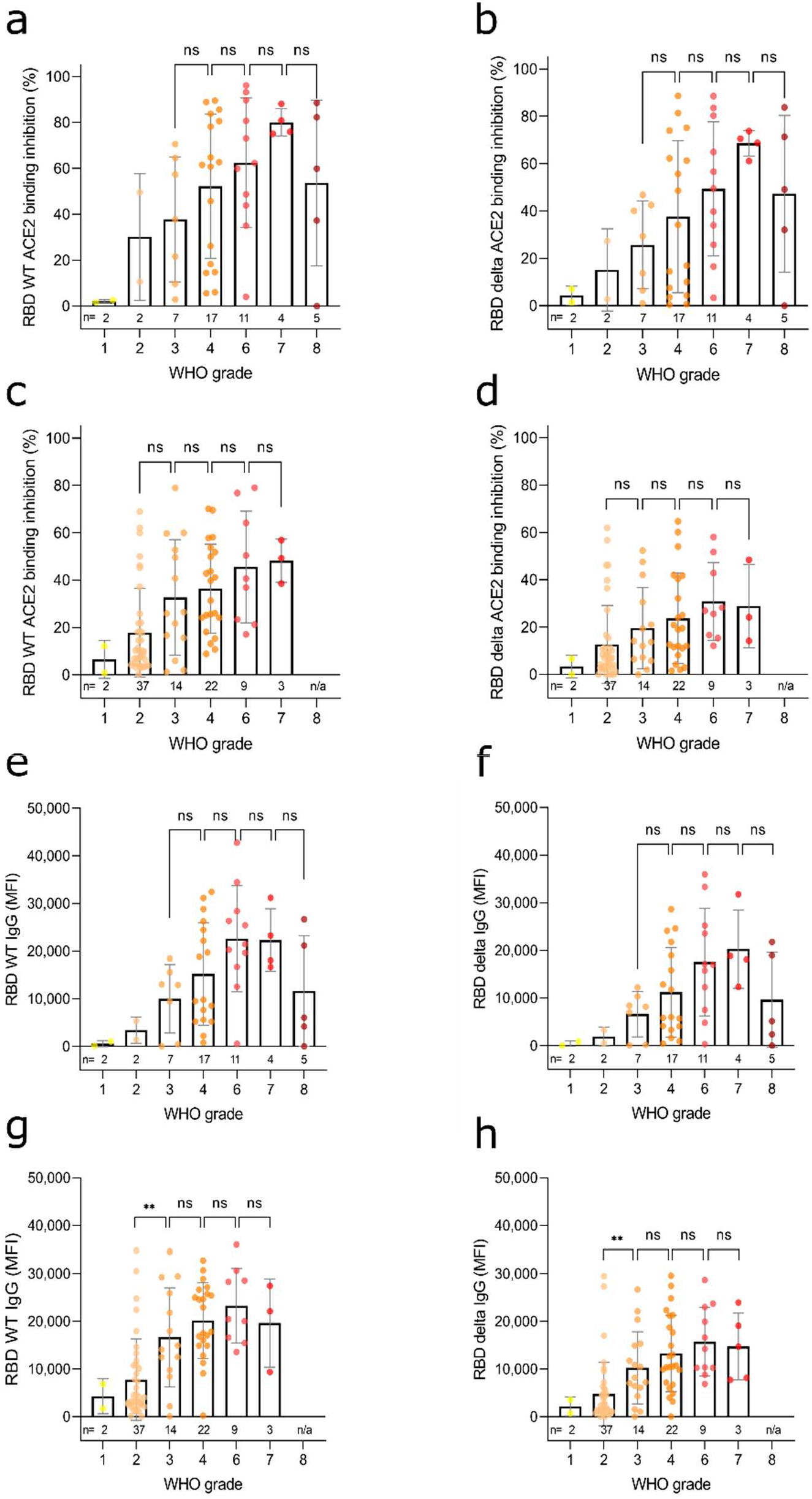
Correlation of anti-RBD IgG levels and ACE2 binding inhibition with SARS-CoV-2 disease severity. Bar charts showing mean ACE2 binding inhibitions (%) against wild-type and RBD delta are correlated with WHO grades for disease severity for samples 7-49 days post PCR (a, b) and ≥ 50 days post PCR (c, d). Mean anti-RBD WT IgG and anti-RBD delta IgG levels are shown for samples 7-49 days post PCR (e, f) and ≥ 50 days post PCR (g, h). Individual samples are displayed as colored dots, bars indicate the mean of the dataset with error bars representing standard deviation. Number of samples is given below the columns (n). If no samples for a group were available, the column is labeled with “n/a”. WHO grade 1 - ambulatory / no limitations of activities, 2 - ambulatory / limitation of activities, 3 - hospitalized, mild disease / no oxygen therapy, 4 - hospitalized, mild disease / mask or nasal prongs, 6 - hospitalized, severe disease / intubation + mechanical ventilation, 7 - hospitalized, severe disease / ventilation + additional organ support (pressors, RRT, ECMO), 8 – Death. The study did not contain samples of WHO grade 5.

## Discussion

With vaccination campaigns now increasingly focusing on the role of booster doses, the quality of immune protection against SARS-CoV-2 in view of constantly emerging variants is of great interest. Whereas in the early-phase of the pandemic SARS-CoV-2 antibody assays were helpful in determining seroprevalence and support vaccine development, now a reliable correlate of immune protection is needed to securely lift social restrictions and guide future vaccine developments.

We show here that the performance of RBDCoV-ACE2 correlates strongly with classical VNTs, confirming that the assay is measuring the activity of neutralizing antibodies, while our technical validation also confirms RBDCoV-ACE2 is stable and reproducible. While cell-culture based VNTs (e.g. plaque reduction neutralization test) are the gold standard for neutralization assays, they have many disadvantages over conventional protein-based surrogate assays. Such assays require rapid access to continually changing virus variants and as such special biosafety level 3 laboratories are necessary. Additionally, VNTs are cell-culture based and therefore it takes multiple days to conduct an experiment with reproducibility potentially affected by either the cells or their long culture conditions. Consequently, highly reproducible assays under substantially faster and safer working conditions (e.g. BSL 1) would be highly beneficial. RBDCoV-ACE2 is finished in under 4 hours and only requires 5 µL of patient sample to measure ACE2 binding inhibition simultaneously against multiple SARS-CoV-2 VOCs and VOIs. As a protein-based assay, it does not require enhanced safety protocols to be followed and can be completed safely in a BSL1 laboratory. Due to the bead-based nature and plate format, it is automatable, suitable for high-throughput, standardized and highly reproducible. The protein-based nature also allows for the rapid inclusion of emerging variants or single mutations. In comparison to the commercially available inhibition assay examined (NeutraLISA), RBDCoV-ACE2 did not have an apparent saturation phase, and therefore has a resolution range that enables greater separation of samples, particularly those which are highly reducing binding inhibition. The stronger correlation of RBDCoV-ACE2 to the VNT compared to NeutraLISA makes it a more accurate alternative to available commercially available inhibition assays.

Similarly to other authors [35,36], we identified a positive correlation between anti-RBD IgG levels and ACE2 binding inhibition, suggesting that neutralizing antibodies represent a consistent portion of all antibodies produced. Similar correlations between anti-S1, anti-trimeric spike and ACE2 binding inhibition as well as no ACE2 binding to the S2 domain percentages reinforce this conclusion. There is however a large degree of individualism in responses, with some samples having low titers yet high ACE2 binding inhibition for specific RBD mutants. We also identified, as other have done previously [6,37], a correlation between disease severity and ACE2 binding inhibition. However, the decrease in IgG levels and ACE2 binding inhibition of patients with WHO disease grade 8 (death) has not to our knowledge been reported before. This decrease requires further investigation to determine its cause given its likely role in patient mortality.

As expected, ACE2 binding inhibition towards VOCs was highly variable. The strongest reductions in binding inhibition were all from variants (eta, gamma, theta and beta) with a E484K mutation. This specific mutation has been reported in multiple studies as an escape mutation that enhances the RBD-ACE2 affinity [38]. ACE2 binding inhibition was further reduced among these variants for those which additionally had a N501Y mutation (gamma, theta and beta), which is known to further enhance RBD-ACE2 binding [39]. These results are in-line with previous findings that have reported significant reductions in neutralization for gamma and beta [40-43]. The gamma and beta RBDs in our assays are only separated by a single K417N mutation, which is known to significantly reduce both the RBD-ACE2 binding affinity as well as the binding affinity to monoclonal therapeutic antibodies or other human antibodies [44]. Among recently emerged strains (delta, kappa, lambda), ACE2 binding inhibition compared to wild-type was reduced for all. The reduction in ACE2 binding inhibition seen for kappa and delta are comparable to recent findings [45], although we could not confirm the reduction seen by other authors for Lambda [46]. This is likely due to the 7-amino acid deletion in the N-terminal domain of lambda’s spike protein, which is not present in the RBD and is thought to contribute to its immune evading properties [47]. Overall the decrease in ACE2 binding inhibition against RBDs of all analyzed variants compared to wild-type has important implications for the design of second generation vaccines.

RBDCoV-ACE2 has limitations similar to other protein-based in vitro neutralization assays, such as only accounting for the Nabs that block the RBD-ACE2 interaction site through steric hindrance, and not for Nabs that interfere with cell entry mechanisms as would be analyzed in a VNT. Furthermore, the binding assay is also more prone to non-specific binding events. However, a major advantage of RBDCoV-ACE2 over VNTs is the speed of response toward viral evolution such as emerging variants of concern. The bead-based format of the assay is also highly reproducible and not susceptible to changes in experimental conditions, as is the case for cell culture based VNTs. The plate format of the assay also enables automation and high-throughput screening. Our assay only requires recombinant expressed RBD proteins which can be quickly and easily produced. Additionally, this assay has the possibility of introducing artificial mutants to screen for possible escape variants that could arise in the future. Among our COVID-19 study population, the majority were admitted to the intensive care unit meaning that the more serious grades of COVID-19 infection are heavily overrepresented in our population, while asymptomatic infections, which are known to be the most common form of disease progression [48], are severely underrepresented. Our sample set for longitudinal analysis is also highly variable in sampling times post-PCR. However, this large variation is also beneficial as it clearly demonstrates the individual variability in ACE2 binding inhibition.

In conclusion, we have developed and validated RBDCoV-ACE2, an ACE2-RBD inhibition assay that analyzes current variants of concern/under investigation/interest of SARS-CoV-2. Assay performance showed good correlation to VNT, confirming that neutralizing antibodies are being analyzed. ACE2 binding inhibition was highly variable among all variants examined, with the 484 aa residue appearing to be critical in reducing ACE2 binding inhibition. ACE2 binding inhibition correlated with both antibody titers and disease severity, although responses were highly individualistic. Overall, the protein-based format of the assay, allows for the fast and simple incorporation of new SARS-CoV-2 variants, enabling rapid screening to identify how ACE2 binding inhibition is altered for emerging variants, or in guiding next-generation vaccine development to target a range of SARS-CoV-2 variants.

## Materials and Methods

### Sample collection for assay validation

16 serum samples consisting of 12 samples from COVID-19 patients (ethical approval #179/2020/BO2, University Hospital Tübingen) and four negative pre-pandemic samples (Central BioHub) were measured by both virus neutralization assay and RBDCoV-ACE2 as part of the assay validation.

For technical assay validation, negative pre-pandemic serum samples were purchased from Central BioHub and four previously collected vaccinated samples from healthcare workers vaccinated with the Pfizer BNT-162b2 vaccine [30] (222/2020/BO2, University Hospital Tübingen) as well as one sample from a COVID-19 infected patient (#179/2020/BO2, University Hospital Tübingen) were used.

### COVID-19 Sample collection

266 serum samples were collected from 168 patients hospitalized at the University Hospital Tübingen, Germany between April 17, 2020 and May 12, 2021. Longitudinal samples were measured from 35 of the 168 patients ranging from 2 to 12 samples per patient. All individuals were tested positive by SARS-CoV-2 PCR. Key characteristics of the study population are summarized in **Table S1**.

For serum collection, blood was extracted by venipuncture, with the serum blood collection tube rotated 180° two to three times to extract possible air bubbles in the sample. After a minimum coagulation time of 30 minutes at room temperature, serum was extracted by centrifugation for 15 minutes at 2000 x g (RT) and then stored at -80 °C until analysis. Time between blood sampling and centrifugation did not exceed 2 hours.

### Ethical Approval

Collection of samples and the execution of this study was approved by the Ethics committee of the Eberhard Karls University Tübingen and the University Hospital Tübingen under the ethical approval numbers 188/2020A and 764/2020/BO2 to Prof. Dr. Michael Bitzer. All participants signed the broad consent of the Medical Faculty Tübingen for sample collection. Samples that were used for assay validation had their collection approved by the Ethics committee of the Eberhard Karls University Tübingen and the University Hospital Tübingen under the ethical approval numbers 222/2020/BO2 to Dr. Karina Althaus and 179/2020/BO2 to Prof. Dr. Juliane Walz. For all assay validation samples, written informed consent was obtained.

### Expression and Purification of SARS-CoV-2 RBD mutants

The expression plasmid pCAGGS, encoding the receptor-binding domain (RBD) of SARS-CoV-2 spike protein (amino acids 319-541), was kindly provided by F. Krammer [49]. Expression and purification of VOCs alpha, beta and epsilon was carried out as previously described [30,50]. RBDs of SARS-CoV-2 VOCs gamma, delta, eta, theta, kappa and A.23.1 were generated by PCR amplification of fragments from wild type or cognate DNA templates and subsequent fusion PCR by overlap extension to introduce described mutations. Based on RBD wild type sequence, primer pairs RBDfor, E484Krev and E484Kfor, RBDrev for VOC eta and RBDfor, V367Frev and V367Ffor, RBDrev for A.23.1 were used. VOC lambda was generated based on RBD wild type sequence using primer pairs L452Qfor, L452Qrev and F490Sfor, F490Srev. VOC delta was generated based on VOC epsilon using primer pairs RBDfor, T478Krev and T478Kfor, RBDrev. Based on VOC alpha sequence, VOC theta was generated using primer pairs RBDfor, E484Krev and E484Kfor, RBDrev. VOC kappa was generated based on VOC eta sequence using primer pairs RBDfor, L452Rrev and L452Rfor, RBDrev. VOC gamma was generated based on VOC theta sequence using primer pairs RBDfor, K417Trev and K417Tfor, RBDrev. Amplificates were inserted into the pCDNA3.4 expression vector using XbaI and NotI restriction sites. The integrity of all expression constructs was confirmed by standard sequencing analysis. An overview of the primer sequences is shown in **Table S2**. Confirmed constructs were expressed in Expi293 cells [30,34]. Briefly, cells were cultivated (37 °C, 125 rpm, 8% (v/v) CO_2_) to a density of 5.5 × 10^6^ cells/mL and diluted with Expi293F expression medium. Transfection of the corresponding plasmids (1 µg/mL) with Expifectamine was performed as per the manufacturer’s instructions. Enhancers were added as per the manufacturer’s instructions 20 h post transfection. Cell suspensions were cultivated for 2–5 days (37 °C, 125 rpm, 8 % (v/v) CO_2_) and centrifuged (4 °C, 23,900×*g*, 20 min) to clarify the supernatant. Afterwards, supernatants were filtered with a 0.22-µm membrane (Millipore, Darmstadt, Germany) and supplemented with His-A buffer stock solution (20 mM Na_2_HPO_4_, 300 mM NaCl, 20 mM imidazole, pH 7.4). The solution was applied to a HisTrap FF crude column on an Äkta pure system (GE Healthcare, Freiburg, Germany), extensively washed with His-A buffer, and eluted with an imidazole gradient (50–400 mM). Amicon 10K centrifugal filter units (Millipore, Darmstadt, Germany) were used for buffer exchange to PBS and concentration of eluted proteins.

### Bead coupling

The in-house expressed RBD mutants were immobilized on magnetic MagPlex beads (Luminex) using the AMG Activation Kit for Multiplex Microspheres (# A-LMPAKMM-400, Anteo Technologies). In brief, 1 mL of spectrally distinct MagPlex beads (1.25 *10^7^ beads) were activated in 1 mL of AnteoBind Activation Reagent for 1 hour at room temperature. The beads were washed twice with 1 mL of conjugation buffer using a magnetic separator, before being resuspended in 1 mL of antigen solution diluted to 25 µg/mL in conjugation buffer. After 1 h incubation at room temperature the beads were washed twice with 1 mL conjugation buffer and incubated for 1 h in 0.1 % (w/v) BSA in conjugation buffer for blocking. Following this, the beads were washed twice with 1 mL storage buffer. Finally, the beads were resuspended in 1 mL storage buffer and stored at 4°C until further use.

### RBDCoV-ACE2

Assay buffer (1:4 Low Cross Buffer (Candor Bioscience GmbH) in CBS (1x PBS + 1% BSA) + 0.05 % Tween20) was supplemented with biotinylated human ACE2 (Sino Biological, # 10108-H08H-B) to a final concentration of 342.9 ng/mL to produce ACE2 buffer. Working inside a sterile laminar flow cabinet, serum samples were thawed and diluted 1:25 in assay buffer before being further diluted 1:8 in ACE2 buffer resulting in a final concentration of 300 ng/mL ACE2 in all 1:200 diluted samples. Spectrally distinct populations of MagPlex beads (Luminex) coupled with RBD proteins of SARS-CoV-2 wild type and variants alpha, beta, gamma, epsilon, eta, theta, kappa, delta, lambda, Cluster 5 and A.23.1 were pooled in assay buffer to create a bead mix (40 beads/µL per bead population). 25 µL of diluted serum was added to 25 µL of bead mix in each well of a 96-well plate (Corning, #3642). To allow comparison of ACE2 binding inhibition between different RBD mutants on a relative scale, 300 ng/mL ACE2 without added serum was measured in triplicates on every plate as normalization control. Additionally, one quality control sample was analyzed in triplicates on every plate. For blank measurement, 25 µL assay buffer instead of diluted sample was added to two wells per plate. Samples were incubated for 2h at 21°C while shaking at 750 rpm on a thermomixer. Following incubation, the beads were washed three times with 100 µL wash buffer (1x PBS + 0.05 Tween20) using a microplate washer (Biotek 405TS, Biotek Instruments GmbH). For detection of bound biotinylated ACE2, 30 µL of 2 µg/mL RPE-Streptavidin was added to each well and the plate was incubated for 45 min at 21 °C while shaking at 750 rpm on a thermomixer. Afterwards, the beads were washed again three times with 100 µL wash buffer. The 96-well plate was placed for 3 min on the thermomixer at 1000 rpm to resuspend the beads before analysis using a FLEXMAP 3D instrument (Luminex) with the following settings: 80 µL (no timeout), 50 events, Gate: 7,500 – 15,000, Reporter Gain: Standard PMT. MFI values of each sample were divided by the mean of the ACE2 normalization control. The normalized values were converted into percent and subtracted from 100 resulting in the percentage of ACE2 binding inhibition. Negative values were manually set to zero.

### MULTICOV-AB

MULTICOV-AB [34], an in-house produced SARS-CoV-2 antibody assay, was performed with all serum samples to measure RBD-specific IgG levels. The antigen panel was expanded to include RBD proteins from 11 different SARS-CoV-2 variants from which all, except the Cluster 5 variant from Sino Biological (# 40592-V08H80), were produced in-house. The assay was carried out as previously described [30].

### Virus Neutralization Assay (VNT)

VNTs for the wild-type (Tü1) variant were performed as previously described [51]. Briefly, 1 × 104 Caco-2 cells/well were seeded in 96-well plates the day before infection in media containing 5% FCS. Caco-2 cells were co-incubated with the SARS-CoV-2 strain icSARS-CoV-2-mNG at a MOI = 1.1 and serum samples in serial dilutions in the indicated concentrations. 48 hpi cells were fixed with 2% PFA and stained with Hoechst33342 (1 µg/mL final concentration) for 10 min at 37°C. The staining solution was removed and exchanged for PBS. For quantification of infection rates, images were taken with the Cytation3 (Biotek Instruments GmbH) and Hoechst+ and mNG+ cells were automatically counted by the Gen5 Software (Biotek Instruments GmbH). Infection rate was determined by dividing the number of infected cells through total cell count per condition. Virus-neutralizing titers (VNT_50_s) were calculated as the half-maximal inhibitory serum dilution.

### Assay validation experiments

To determine the intra-assay precision of RBDCoV-ACE2, 12 replicates of four serum samples (Vac1 – Vac4) were measured on a 96-well plate (Corning, #3642). Additionally, 15 replicates of the 300 ng/mL ACE2 control and 12 replicates of the blank control containing only assay buffer without sample or ACE2 were measured. For inter-assay precision, five serum samples (Vac1 – Vac4 and Inf1) were measured in triplicates in four independent experiments. Additionally, the quality control, the ACE2 normalization control and blank were also processed in triplicates in the same four experiments. Short-term stability was determined by storing ACE2 buffer under six different conditions before proceeding with the assay protocol. The prepared ACE2 buffer was stored 2 h, 4 h and 24 h at both 4°C and room temperature and compared to ACE2 buffer without storage (fresh). Replicate MFI values of every sample (Vac1 – Vac4 (vaccinated), Inf1 (infected) and pre-pandemic) were normalized to the values of the respective normalization ACE2 control. Freeze-thaw stability of the biotinylated ACE2 stocks was determined by analyzing six serum samples (Vac1 – Vac4 (vaccinated), Inf1 (infected) and pre-pandemic) in triplicates, with ACE2 stocks undergoing 1 to 5 cycles. In addition to that, every sample was also processed with ACE2 not re-frozen once thawed (fresh, 0 freeze-thaw cycles). The MFI values of every sample were normalized to the values of the respective ACE2 normalization control. To investigate the stability of RBDCoV-ACE2 against variations of the used ACE2 concentration, six samples (Vac1 – Vac4 (vaccinated), Inf1 (infected) and pre-pandemic) were analyzed with ACE2 concentrations ranging from 150 ng/mL to 350 ng/mL. Replicate MFI values of every sample were normalized to the values of the respective ACE2 normalization control. For analysis, the mean, standard deviation and coefficient of variation in percent of all replicates were calculated.

To confirm, that the multiplex assay format has no undesirable effect on ACE2 binding inhibition values compared to singleplex measurements, 24 samples (pre-pandemic (n=5) and COVID-19 infected (n=19)) were analyzed in both singleplex and multiplex (for all VOCs).

### NeutraLISA

One sample from each individual donor (n=168) was analyzed with the commercially available in-vitro diagnostic test SARS-CoV-2 NeutraLISA (Euroimmun). The assay was performed according to the manufacturer’s instructions. For longitudinal donors with more than one sample available, the sample closest to 20 days after positive PCR diagnosis was picked. Negative values were manually set to zero.

### Statistical analysis

Data collection and assignment to metadata was performed with Microsoft Excel 2016. Data analysis, visualization and curve fitting was performed with Graphpad Prism (version 9.1.2). Virus-neutralizing titers (VNT_50_s) as the half-maximal inhibitory serum dilution were calculated using 4-parameter nonlinear regression. Longitudinal curves were fitted using a one-site total binding equation. Correlations were analyzed using Spearman’s correlation coefficient. Significances were calculated (where appropriate) using Mann-Whitney U tests. Figures were edited with Inkscape (version 0.92.4). Data generated for this manuscript is available from the authors upon request.

## Data Availability

All data generated in this manuscript is available from the authors upon request.

## Acknowledgements

We thank Johanna Griesbaum, Jennifer Jüngling and Christine Geisler for excellent technical assistance. This work was financially supported by the State Ministry of Baden-Württemberg for Economic Affairs, Labour and Housing Construction (grant number FKZ 3-4332.62-NMI-68). The funders had no role in study design, data collection and interpretation, or the decision to submit the work for publication.

## Author contributions

DJ and AD conceived the study and designed the experiments. NSM supervised the study. DJ and MaB performed the experiments. DJ, AD, MaB produced components for RBDCoV-ACE2. KK, SB, CS, HH, KS, NM, KA, MK, JW, MB and SG collected samples or organized their collection and provided clinical metadata. NR and MS provided virus neutralization test data. PDK, BT, TW and UR produced and designed recombinant assay proteins. MB, SG and NSM procured funding. DJ performed data analysis and generated the figures. DJ and AD wrote the manuscript. All authors critically reviewed and approved the final manuscript.

## Supplementary Material Legends

Table S1 | Characteristics of the analyzed COVID-19 serum sample collection set.

Table S2 | Primer sequences used for expression of RBD mutants.

Table S3 | RBDCoV-ACE2 technical validation results. Percentage coefficients of variation (%CV) of normalized MFI values for every SARS-CoV-2 RBD for all analyzed samples (n=6) including the mean of all samples.

Figure S1 | RBDCoV-ACE2 technical validation results. Results of intra-assay precision (a), inter-assay precision (b), short-term stability (c), freeze-thaw stability (d) and parallelism (e) experiments analyzing ACE2 binding inhibition (displayed as %) using wild-type (WT) RBD. Four samples from donors vaccinated with Pfizer BNT-162b2 (n=4), one COVID-19 infected (n=1) and one pre-pandemic sample (n=1, grey) were analyzed. Data points of each sample are illustrated by different shapes according to the figure key. Percent coefficients of variation (%CV) for all included RBD mutants are summarized in Table S3.

Figure S2 | Comparison of multiplex and singleplex assay formats. Linear regression analysis between ACE2 binding inhibition (%) values of samples from pre-pandemic (n=5) and COVID-19 infected (n=19) individuals analyzed in both multiplex and singleplex for RBD WT (a) and RBD delta (b). Correlation analysis was performed after Spearman and the correlation coefficient r is shown.

Figure S3 | Correlation between IgG MFI signals and ACE2 binding inhibition (%) against SARS-CoV-2 S1-domain (a) and trimeric spike (b) of serum samples from COVID-19 patients (n= 168). Regression analysis comparing ACE2 binding inhibitions (%) for S1 and trimeric spike with RBDCoV-ACE2 results of RBD WT (c and d). Each circle represents one sample (n=168). For longitudinal donors with more than one sample available, the sample closest to 20 days post positive PCR diagnosis was selected. Spearman’s correlation coefficient (r) as well as the equation of the linear regression is specified for every correlation.

Figure S4 | Relation between ACE2 binding inhibition (%) and gender, donor age and Body-mass-index (BMI). Correlation between wild-type ACE2 binding inhibition (%) and gender (a, b), age (c, d) and BMI (e, f) for samples 7-49 days post PCR (a, c, e) and ≥ 50 days post PCR (b, d, f). P-values, when significant, are shown for all panels. Spearman’s r was used to determine correlations.

## References

1 Forthal, D. N. Functions of Antibodies. Microbiology spectrum 2, Aid-0019-2014, doi:10.1128/microbiolspec.AID-0019-2014 (2014).

2 Jiang, S., Zhang, X., Yang, Y., Hotez, P. J. & Du, L. Neutralizing antibodies for the treatment of COVID-19. Nature biomedical engineering 4, 1134–1139, doi:10.1038/s41551-020-00660-2 (2020).

3 Shi, R. et al. A human neutralizing antibody targets the receptor-binding site of SARS-CoV-2. Nature 584, 120–124, doi:10.1038/s41586-020-2381-y (2020).

4 Piccoli, L. et al. Mapping Neutralizing and Immunodominant Sites on the SARS-CoV-2 Spike Receptor-Binding Domain by Structure-Guided High-Resolution Serology. Cell 183, 1024–1042.e1021, doi:10.1016/j.cell.2020.09.037 (2020).

5 Dispinseri, S. et al. Neutralizing antibody responses to SARS-CoV-2 in symptomatic COVID-19 is persistent and critical for survival. Nature communications 12, 2670, doi:10.1038/s41467-021-22958-8 (2021).

6 Garcia-Beltran, W. F. et al. COVID-19-neutralizing antibodies predict disease severity and survival. Cell 184, 476–488.e411, doi:10.1016/j.cell.2020.12.015 (2021).

7 McMahan, K. et al. Correlates of protection against SARS-CoV-2 in rhesus macaques. Nature 590, 630–634, doi:10.1038/s41586-020-03041-6 (2021).

8 Kim, Y. I. et al. Critical role of neutralizing antibody for SARS-CoV-2 reinfection and transmission. Emerging microbes & infections 10, 152–160, doi:10.1080/22221751.2021.1872352 (2021).

9 Rogers, T. F. et al. Isolation of potent SARS-CoV-2 neutralizing antibodies and protection from disease in a small animal model. Science (New York, N.Y.) 369, 956–963, doi:10.1126/science.abc7520 (2020).

10 Weinreich, D. M. et al. REGN-COV2, a Neutralizing Antibody Cocktail, in Outpatients with Covid-19. The New England journal of medicine 384, 238–251, doi:10.1056/NEJMoa2035002 (2021).

11 (FDA), F. a. D. A. Coronavirus (COVID-19) Update: FDA Authorizes Monoclonal Antibodies for Treatment of COVID-19, <https://www.fda.gov/news-events/press-announcements/coronavirus-covid-19-update-fda-authorizes-monoclonal-antibodies-treatment-covid-19> (2020).

12 Gottlieb, R. L. et al. Effect of Bamlanivimab as Monotherapy or in Combination With Etesevimab on Viral Load in Patients With Mild to Moderate COVID-19: A Randomized Clinical Trial. Jama 325, 632–644, doi:10.1001/jama.2021.0202 (2021).

13 (FDA), F. a. D. A. Coronavirus (COVID-19) Update: FDA Authorizes Monoclonal Antibodies for Treatment of COVID-19, <https://www.fda.gov/news-events/press-announcements/coronavirus-covid-19-update-fda-authorizes-monoclonal-antibodies-treatment-covid-19-0> (2021).

14 Zhou, P. et al. A pneumonia outbreak associated with a new coronavirus of probable bat origin. Nature, 1–4 (2020).

15 Graham, M. S. et al. Changes in symptomatology, reinfection, and transmissibility associated with the SARS-CoV-2 variant B.1.1.7: an ecological study. The Lancet. Public health 6, e335–e345, doi:10.1016/s2468-2667(21)00055-4 (2021).

16 Tegally, H. et al. Detection of a SARS-CoV-2 variant of concern in South Africa. Nature 592, 438–443, doi:10.1038/s41586-021-03402-9 (2021).

17 Sabino, E. C. et al. Resurgence of COVID-19 in Manaus, Brazil, despite high seroprevalence. Lancet (London, England) 397, 452–455, doi:10.1016/s0140-6736(21)00183-5 (2021).

18 Control, E. C. f. D. P. a. Threat Assessment Brief: Emergence of SARS-CoV-2 B.1.617 variants in India and situation in the EU/EEA, <https://www.ecdc.europa.eu/en/publications-data/threat-assessment-emergence-sars-cov-2-b1617-variants> (2021).

19 (WHO), W. H. O. (2021).

20 (WHO), W. H. O. Tracking SARS-CoV-2 variants, <https://www.who.int/en/activities/tracking-SARS-CoV-2-variants/> (2021).

21 O’Toole, A. & Hill, V. Lineage C.37, <https://cov-lineages.org/lineage.htmlãlineage=C.37> (2021).

22 Skowronski, D. M. & De Serres, G. Safety and Efficacy of the BNT162b2 mRNA Covid-19 Vaccine. The New England journal of medicine 384, 1576–1577, doi:10.1056/NEJMc2036242 (2021).

23 Baden, L. R. et al. Efficacy and Safety of the mRNA-1273 SARS-CoV-2 Vaccine. The New England journal of medicine 384, 403–416, doi:10.1056/NEJMoa2035389 (2021).

24 Voysey, M. et al. Safety and efficacy of the ChAdOx1 nCoV-19 vaccine (AZD1222) against SARS-CoV-2: an interim analysis of four randomised controlled trials in Brazil, South Africa, and the UK. Lancet (London, England) 397, 99–111, doi:10.1016/s0140-6736(20)32661-1 (2021).

25 Sadoff, J. et al. Safety and Efficacy of Single-Dose Ad26.COV2.S Vaccine against Covid-19. The New England journal of medicine 384, 2187–2201, doi:10.1056/NEJMoa2101544 (2021).

26 Krammer, F. SARS-CoV-2 vaccines in development. Nature 586, 516–527, doi:10.1038/s41586-020-2798-3 (2020).

27 Stamatatos, L. et al. mRNA vaccination boosts cross-variant neutralizing antibodies elicited by SARS-CoV-2 infection. Science (New York, N.Y.), doi:10.1126/science.abg9175 (2021).

28 Jalkanen, P. et al. COVID-19 mRNA vaccine induced antibody responses against three SARS-CoV-2 variants. Nature communications 12, 3991, doi:10.1038/s41467-021-24285-4 (2021).

29 Shen, X. et al. Neutralization of SARS-CoV-2 Variants B.1.429 and B.1.351. The New England journal of medicine 384, 2352–2354, doi:10.1056/NEJMc2103740 (2021).

30 Becker, M. et al. Immune response to SARS-CoV-2 variants of concern in vaccinated individuals. Nature communications 12, 3109, doi:10.1038/s41467-021-23473-6 (2021).

31 Hoffmann, M. et al. SARS-CoV-2 variants B.1.351 and P.1 escape from neutralizing antibodies. Cell 184, 2384–2393.e2312, doi:10.1016/j.cell.2021.03.036 (2021).

32 Krammer, F. A correlate of protection for SARS-CoV-2 vaccines is urgently needed. Nature medicine, doi:10.1038/s41591-021-01432-4 (2021).

33 Euroimmun. SARS-CoV-2 NeutraLISA: Produkt-Datenblatt, <https://www.coronavirus-diagnostik.de/documents/Indications/Infections/Coronavirus/EI_2606_D_DE_F.pdf> (2021).

34 Becker, M. et al. Exploring beyond clinical routine SARS-CoV-2 serology using MultiCoV-Ab to evaluate endemic coronavirus cross-reactivity. Nature communications 12, 1152, doi:10.1038/s41467-021-20973-3 (2021).

35 Mendrone-Junior, A. et al. Correlation between SARS-COV-2 antibody screening by immunoassay and neutralizing antibody testing. Transfusion 61, 1181–1190, doi:10.1111/trf.16268 (2021).

36 Grenache, D. G., Ye, C. & Bradfute, S. B. Correlation of SARS-CoV-2 Neutralizing Antibodies to an Automated Chemiluminescent Serological Immunoassay. The journal of applied laboratory medicine 6, 491–495, doi:10.1093/jalm/jfaa195 (2021).

37 Legros, V. et al. A longitudinal study of SARS-CoV-2-infected patients reveals a high correlation between neutralizing antibodies and COVID-19 severity. Cellular & molecular immunology 18, 318–327, doi:10.1038/s41423-020-00588-2 (2021).

38 Nelson, G. et al. Molecular dynamic simulation reveals E484K mutation enhances spike RBD-ACE2 affinity and the combination of E484K, K417N and N501Y mutations (501Y.V2 variant) induces conformational change greater than N501Y mutant alone, potentially resulting in an escape mutant. bioRxiv,2021.2001.2013.426558, doi:10.1101/2021.01.13.426558 (2021).

39 Luan, B., Wang, H. & Huynh, T. Enhanced binding of the N501Y-mutated SARS-CoV-2 spike protein to the human ACE2 receptor: insights from molecular dynamics simulations. FEBS letters 595, 1454–1461, doi:10.1002/1873-3468.14076 (2021).

40 Shen, X. et al. SARS-CoV-2 variant B.1.1.7 is susceptible to neutralizing antibodies elicited by ancestral spike vaccines. Cell host & microbe 29, 529–539.e523, doi:10.1016/j.chom.2021.03.002 (2021).

41 Wall, E. C. et al. Neutralising antibody activity against SARS-CoV-2 VOCs B.1.617.2 and B.1.351 by BNT162b2 vaccination. Lancet (London, England) 397, 2331–2333, doi:10.1016/s0140-6736(21)01290-3 (2021).

42 Wang, P. et al. Antibody resistance of SARS-CoV-2 variants B.1.351 and B.1.1.7. Nature 593, 130–135, doi:10.1038/s41586-021-03398-2 (2021).

43 Heggestad, J. T. et al. Rapid test to assess the escape of SARS-CoV-2 variants of concern. Science advances 7, eabl7682, doi:10.1126/sciadv.abl7682 (2021).

44 Luan, B. & Huynh, T. Insights into SARS-CoV-2’s Mutations for Evading Human Antibodies: Sacrifice and Survival. Journal of medicinal chemistry, doi:10.1021/acs.jmedchem.1c00311 (2021).

45 Edara, V. V. et al. Infection and Vaccine-Induced Neutralizing-Antibody Responses to the SARS-CoV-2 B.1.617 Variants. The New England journal of medicine 385, 664–666, doi:10.1056/NEJMc2107799 (2021).

46 Kimura, I. et al. SARS-CoV-2 Lambda variant exhibits higher infectivity and immune resistance. bioRxiv, 2021.2007.2028.454085, doi:10.1101/2021.07.28.454085 (2021).

47 Acevedo, M. L. et al. Infectivity and immune escape of the new SARS-CoV-2 variant of interest Lambda. medRxiv, 2021.2006.2028.21259673, doi:10.1101/2021.06.28.21259673 (2021).

48 Subramanian, R., He, Q. & Pascual, M. Quantifying asymptomatic infection and transmission of COVID-19 in New York City using observed cases, serology, and testing capacity. Proceedings of the National Academy of Sciences of the United States of America 118, doi:10.1073/pnas.2019716118 (2021).

49 Amanat, F. et al. A serological assay to detect SARS-CoV-2 seroconversion in humans. Nature medicine 26, 1033–1036, doi:10.1038/s41591-020-0913-5 (2020).

50 Wagner, T. R. et al. A broadly neutralizing biparatopic Nanobody protects mice from lethal challenge with SARS-CoV-2 variants of concern. bioRxiv, 2021.2008.2008.455562, doi:10.1101/2021.08.08.455562 (2021).

51 Ruetalo, N. et al. Antibody Response against SARS-CoV-2 and Seasonal Coronaviruses in Nonhospitalized COVID-19 Patients. mSphere 6, doi:10.1128/mSphere.01145-20 (2021).

