## Supplementary Material for "COVID-19 patient serum less potently inhibits ACE2-RBD binding for various SARS-CoV-2 RBD mutants"

| ^1^ | NMI Natural and Medical Sciences Institute at the University of Tübingen, Reutlingen, Germany |
| --- | --- |
| ^2^ | Pharmaceutical Biotechnology, Eberhard Karls University, Tübingen, Germany |
| ^3^ | Department Internal Medicine I, University Hospital Tübingen, Tübingen, Germany |
| ^4^ | Institute for Medical Microbiology and Hygiene, University Hospital Tübingen, Tübingen, Germany |
| ^5^ | German Centre for Infection Research (DZIF), Partner Site Tübingen, Tübingen, Germany |
| ^6^ | Institute for Medical Virology and Epidemiology, University Hospital Tübingen, Tübingen, Germany |
| ^7^ | Center for Personalized Medicine, Eberhard Karls University, Tübingen, Germany |
| ^8^ | Institute for Clinical and Experimental Transfusion Medicine, University Hospital Tübingen, Tübingen, Germany |
| ^9^ | Department of Anesthesiology and Intensive Care Medicine, University Hospital Tübingen, Tübingen, Germany |
| ^10^ | Clinical Collaboration Unit Translational Immunology, German Cancer Consortium (DKTK), Department of Internal Medicine, University Hospital Tübingen, Tübingen, Germany |
| ^11^ | Institute for Cell Biology, Department of Immunology, University of Tübingen, Tübingen, Germany |
| ^12^ | Cluster of Excellence iFIT (EXC2180) “Image-Guided and Functionally Instructed Tumor Therapies”, University of Tübingen, Tübingen, Germany |
| ^13^ | Dr. Margarete Fischer-Bosch-Institute for Clinical Pharmacology and Robert Bosch Center for Tumor Diseases (RBCT), Stuttgart, Germany |

Running Head: Less potent ACE2 binding inhibition against SARS-CoV-2 RBD mutants

* denotes shared senior authorship and corresponding authors.

Contact Information

Table S1 | Characteristics of the analyzed COVID-19 serum sample collection set.

| Characteristic |  |
| --- | --- |
| Number of donors | 168 |
| Number of samples | 266 |
| Median age (IQR) -years | 62 (23) |
| Female sex (%) | 78 (46.4) |
| Median ΔT post first positive PCR test (range) | 91 (1-348) |
| Median BMI (range) | 26.9 (18.4 – 50.2) |

Table S2 | Primer sequences used for expression of RBD mutants.

| Primer Name | Sequence (5‘ to 3‘) |
| --- | --- |
| RBDfor | ATATCTAGAGCCACCATGTTCGTGTTTCTGG |
| E484Krev | GCAGTTGAAGCCTTTCACGCCGTTACAAGGGGT |
| E484Kfor | GTAACGGCGTGAAAGGCTTCAACTGCTACTTCCC |
| RBD rev | AAGATCTGCTAGCTCGAGTCGC |
| V367Frev | CGGAGTTGTACAGGAAGGAGTAGTCGGCCACGCA |
| V367Ffor | CGACTACTCCTTCCTGTACAACTCCGCCAGCTTC |
| L452Qfor | GGCAACTACAATTACCAGTACCGGCTGTTCCGGAAG |
| L452Qrev | CGGTACTGGTAATTGTAGTTGCCGCCG |
| F490Sfor | TCAACTGCTACTCCCCACTGCAGTCCTACGGC |
| F490Srev | CTGCAGTGGGGAGTAGCAGTTGAAGCCTTCCAC |
| T478Krev | CGTTACAAGGCTTGCTGCCGGCCTGATAGA |
| T478Kfor | CCGGCAGCAAGCCTTGTAACGGCGTGGAAG |
| L452Rrev | CGGTACCGGTAATTGTAGTTGCCGCCG |
| L452Rfor | GGCAACTACAATTACCGGTACCGGCTGTTCCGGAAG |
| K417Trev | GTTGTAGTCGGCGATGGTGCCTGTCTGTCCAGGGG) |
| K417Tfor | GACAGACAGGCACCATCGCCGACTACAACTACAAG |

Table S3 | RBDCoV-ACE2 technical validation results. Percentage coefficients of variation (%CV) of normalized MFI values for every SARS-CoV-2 RBD for all analyzed samples (n=6) including the mean of all samples.

|  | **Samples** | **RBD WT** | **alpha** | **beta** | **gamma** | **epsilon** | **Cluster 5** | **A.23.1** | **eta** | **theta** | **kappa** |
| --- | --- | --- | --- | --- | --- | --- | --- | --- | --- | --- | --- |
| **Intra-assay precision (%CV)** | Vac1 | 1.4 | 1.5 | 2.6 | 2.5 | 1.3 | 1.5 | 1.7 | 1.4 | 2.1 | 1.1 |
|  | Vac2 | 2.2 | 2.1 | 1.1 | 1.6 | 1.7 | 1.2 | 1.5 | 1.9 | 1.6 | 2.0 |
|  | Vac3 | 2.8 | 4.0 | 3.8 | 2.1 | 4.2 | 3.0 | 3.5 | 2.1 | 1.6 | 4.3 |
|  | Vac4 | 2.2 | 2.1 | 3.4 | 1.9 | 2.0 | 2.0 | 2.3 | 1.7 | 2.1 | 2.4 |
|  | **Mean** | **2.2** | **2.4** | **2.7** | **2.0** | **2.3** | **1.9** | **2.3** | **1.8** | **1.9** | **2.5** |
| **Inter-assay precision (%CV)** | Vac1 | 1.9 | 2.7 | 3.0 | 2.9 | 2.8 | 2.3 | 3.5 | 2.7 | 2.9 | 2.9 |
|  | Vac2 | 2.6 | 3.0 | 3.9 | 2.8 | 3.5 | 2.4 | 2.7 | 3.0 | 2.1 | 3.0 |
|  | Vac3 | 5.7 | 6.9 | 6.9 | 3.5 | 6.4 | 4.6 | 5.5 | 4.9 | 3.6 | 5.2 |
|  | Vac4 | 4.4 | 3.5 | 4.6 | 2.9 | 4.3 | 2.5 | 1.9 | 3.0 | 2.7 | 3.3 |
|  | Inf1 | 1.9 | 2.5 | 2.9 | 2.9 | 2.8 | 2.4 | 1.6 | 1.8 | 1.8 | 2.4 |
|  | **Mean** | **3.3** | **3.7** | **4.3** | **3.0** | **4.0** | **2.8** | **3.0** | **3.1** | **2.6** | **3.4** |
| **Short-term stability (%CV)** | Vac1 | 2.5 | 3.9 | 3.2 | 2.3 | 2.5 | 2.3 | 2.6 | 2.1 | 2.8 | 3.2 |
|  | Vac2 | 2.3 | 3.1 | 4.5 | 2.3 | 2.3 | 2.8 | 2.6 | 2.0 | 2.4 | 2.6 |
|  | Vac3 | 8.6 | 9.3 | 6.4 | 3.0 | 6.9 | 4.4 | 4.9 | 7.1 | 3.2 | 7.3 |
|  | Vac4 | 11.6 | 4.9 | 3.4 | 3.2 | 3.0 | 2.4 | 3.1 | 3.2 | 2.3 | 3.3 |
|  | Inf1 | 1.4 | 2.6 | 3.5 | 2.5 | 1.9 | 2.4 | 2.2 | 1.8 | 1.7 | 1.7 |
|  | Pre-pandemic | 1.6 | 2.3 | 3.7 | 1.9 | 1.9 | 1.8 | 2.2 | 1.8 | 2.4 | 2.1 |
|  | **Mean** | **4.7** | **4.4** | **4.1** | **2.5** | **3.1** | **2.7** | **2.9** | **3** | **2.5** | **3.4** |
| **Freeze-thaw stability (%CV)** | Vac1 | 2.7 | 3.8 | 4.5 | 3.1 | 4.0 | 2.7 | 3.3 | 2.1 | 2.9 | 3.2 |
|  | Vac2 | 4.0 | 3.2 | 2.5 | 2.5 | 3.8 | 2.8 | 2.9 | 2.6 | 3.2 | 2.9 |
|  | Vac3 | 9.3 | 10.2 | 4.7 | 5.8 | 12.1 | 8.9 | 7.3 | 9.8 | 5.5 | 10.6 |
|  | Vac4 | 7.6 | 6.2 | 6.1 | 5.3 | 7.2 | 5.8 | 6.6 | 6.3 | 4.9 | 6.3 |
|  | Inf1 | 3.6 | 2.6 | 4.1 | 3.1 | 3.1 | 2.9 | 2.4 | 2.5 | 2.4 | 3.5 |
|  | Pre-pandemic | 1.8 | 1.8 | 3.2 | 3.7 | 1.8 | 1.4 | 2.1 | 2.3 | 2.3 | 2.0 |
|  | **Mean** | **4.8** | **4.6** | **4.2** | **3.9** | **5.3** | **4.1** | **4.1** | **4.3** | **3.5** | **4.8** |
| **Parallelism (%CV)** | Vac1 | 4.1 | 3.9 | 5.0 | 4.0 | 4.5 | 4.2 | 4.2 | 3.2 | 2.6 | 3.9 |
|  | Vac2 | 4.2 | 2.5 | 4.0 | 3.4 | 2.7 | 2.5 | 4.2 | 2.6 | 2.8 | 4.2 |
|  | Vac3 | 12.0 | 11.7 | 4.0 | 5.3 | 11.9 | 11.8 | 9.3 | 13.6 | 4.5 | 12.7 |
|  | Vac4 | 11.3 | 9.6 | 6.5 | 6.9 | 10.0 | 8.7 | 8.6 | 9.9 | 4.4 | 9.3 |
|  | Inf1 | 5.2 | 3.4 | 5.5 | 3.4 | 4.8 | 4.1 | 4.0 | 4.2 | 4.2 | 3.6 |
|  | Pre-pandemic | 2.1 | 1.2 | 3.6 | 2.8 | 2.3 | 2.1 | 2.4 | 1.9 | 2.7 | 1.4 |
|  | **Mean** | **6.5** | **5.4** | **4.8** | **4.3** | **6.0** | **5.6** | **5.5** | **5.9** | **3.5** | **5.9** |

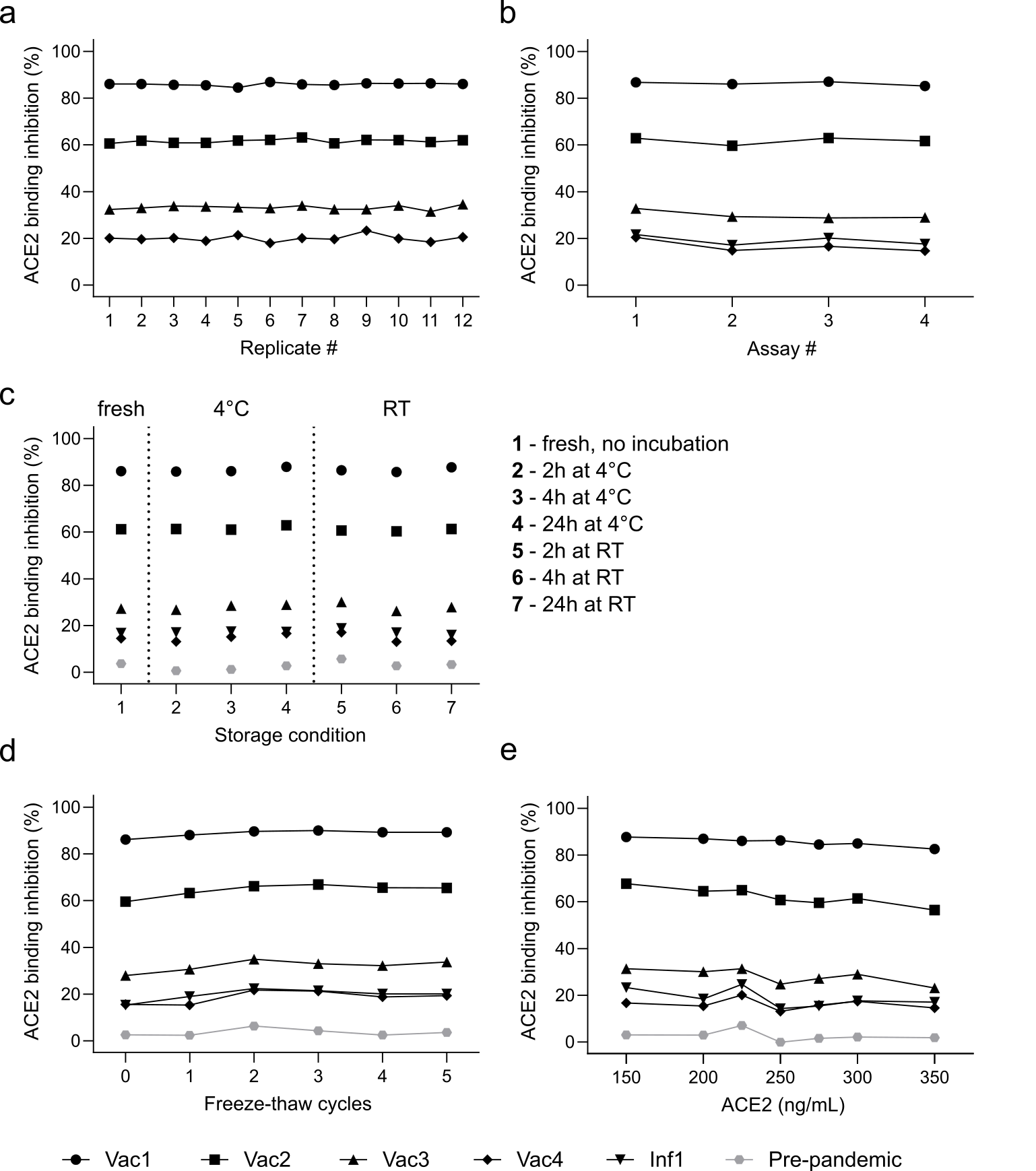

Figure S1 | RBDCoV-ACE2 technical validation results. Results of intra-assay precision (a), inter-assay precision (b), short-term stability (c), freeze-thaw stability (d) and parallelism (e) experiments analyzing ACE2 binding inhibition (displayed as %) using wild-type (WT) RBD. Four samples from donors vaccinated with Pfizer BNT-162b2 (n=4), one COVID-19 infected (n=1) and one pre-pandemic sample (n=1, grey) were analyzed. Data points of each sample are illustrated by different shapes according to the figure key. Percent coefficients of variation (%CV) for all included RBD mutants are summarized in **Table S3**.

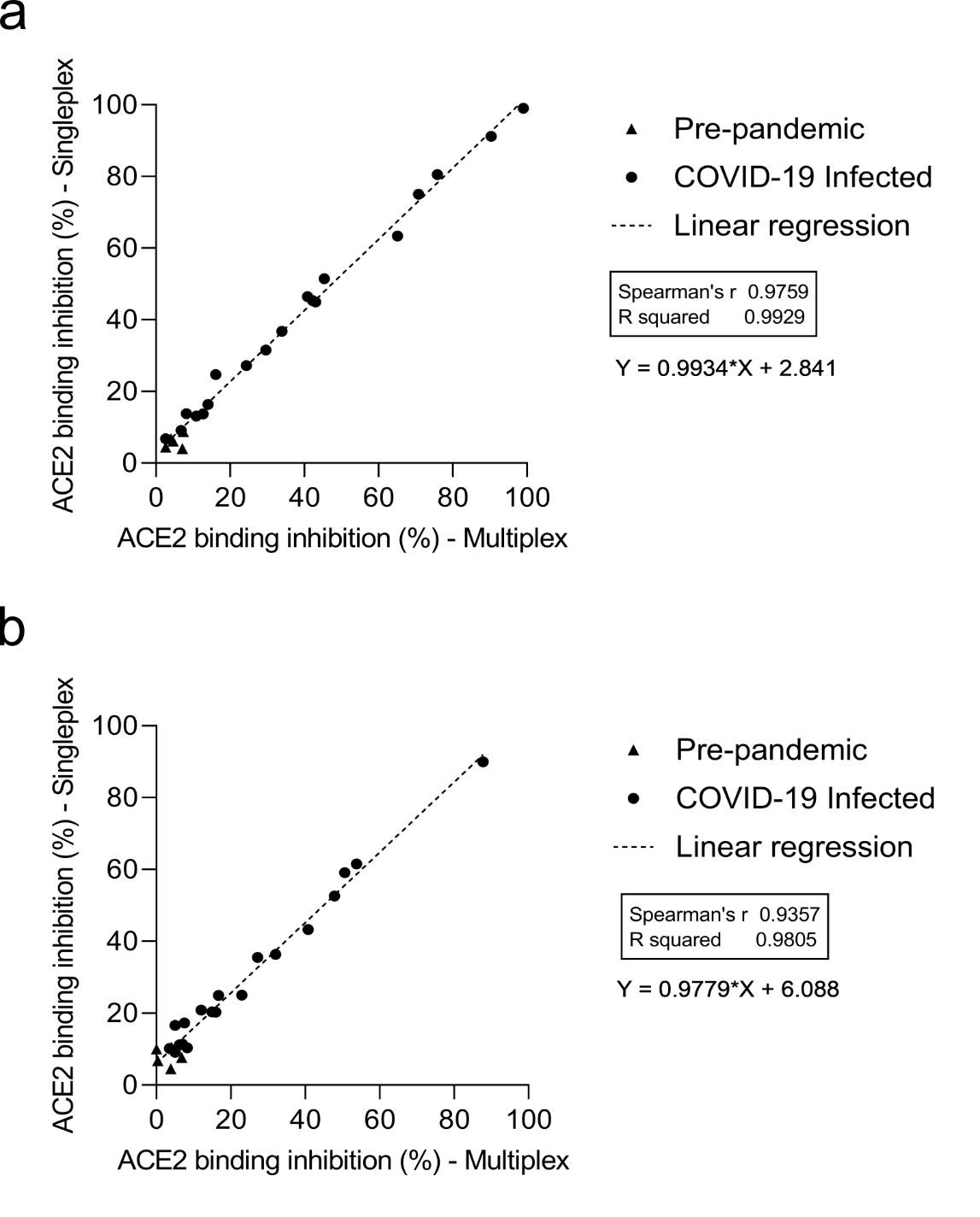

Figure S2 | Comparison of multiplex and singleplex assay formats. Linear regression analysis between ACE2 binding inhibition (%) values of samples from pre-pandemic (n=5) and COVID-19 infected (n=19) individuals analyzed in both multiplex and singleplex for RBD WT (a) and RBD delta (b). Correlation analysis was performed after Spearman and the correlation coefficient r is shown.

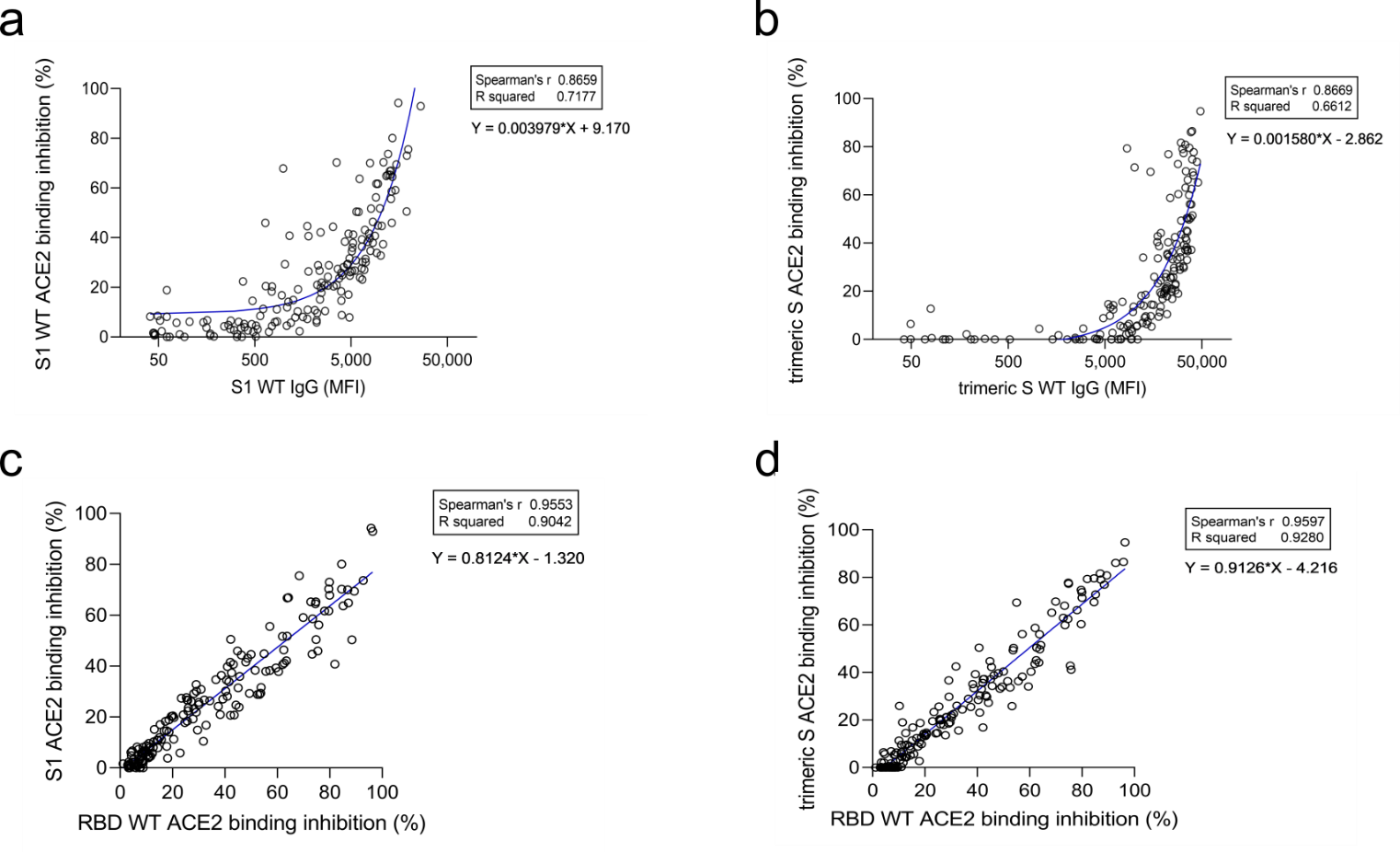

Figure S3 | Correlation between IgG MFI signals and ACE2 binding inhibition (%) against SARS-CoV-2 S1-domain (a) and trimeric spike (b) of serum samples from COVID-19 patients (n= 168). Regression analysis comparing ACE2 binding inhibitions (%) for S1 and trimeric spike with RBDCoV-ACE2 results of RBD WT (c and d). Each circle represents one sample (n=168). For longitudinal donors with more than one sample available, the sample closest to 20 days post positive PCR diagnosis was selected. Spearman’s correlation coefficient (r) as well as the equation of the linear regression is specified for every correlation.

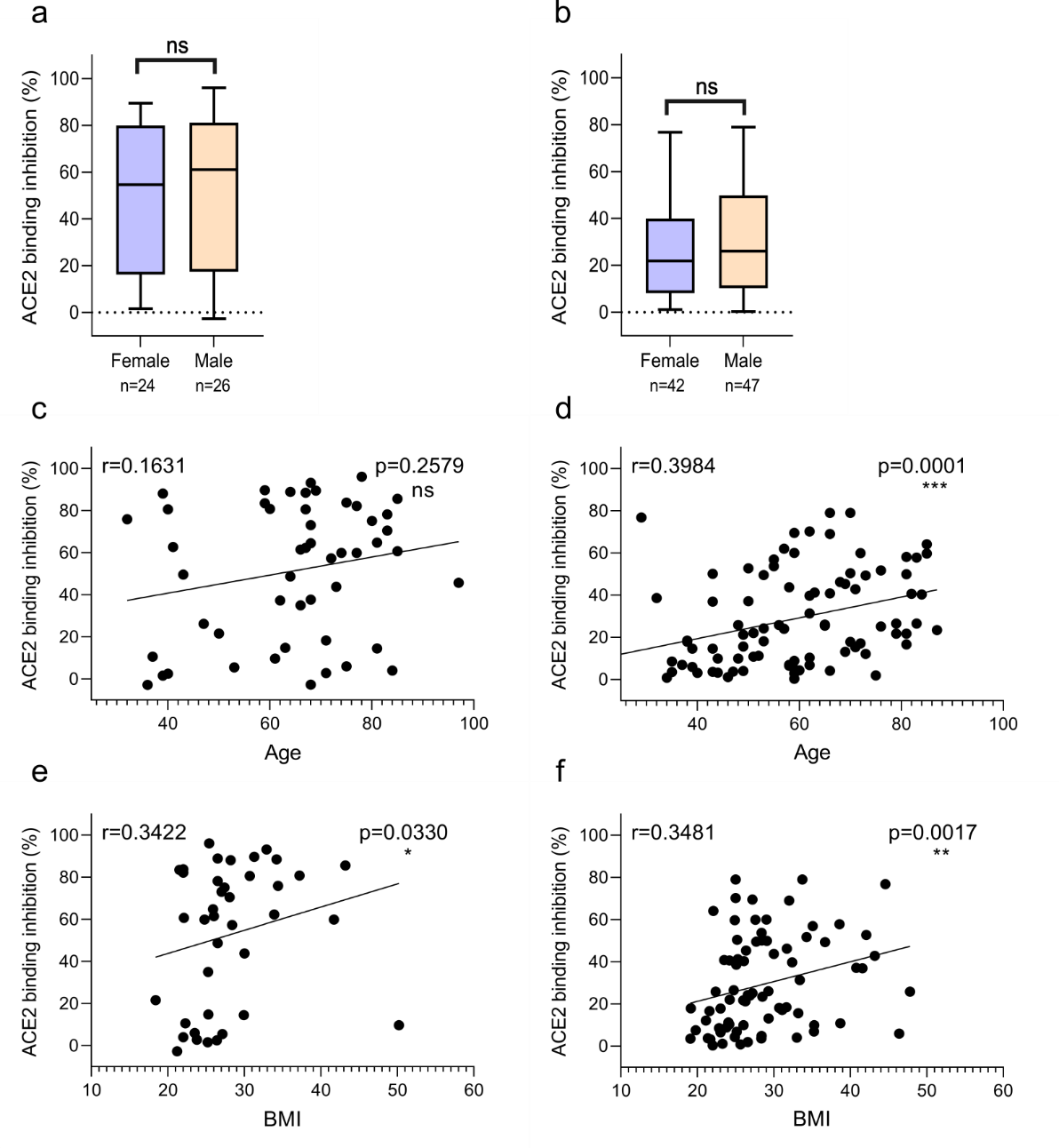

Figure S4 | Relation between ACE2 binding inhibition (%) and gender, donor age and Body-mass-index (BMI). Correlation between wild-type ACE2 binding inhibition (%) and gender (a, b), age (c, d) and BMI (e, f) for samples 7-49 days post PCR (a, c, e) and ≥ 50 days post PCR (b, d, f). P-values, when significant, are shown for all panels. Spearman’s r was used to determine correlations.
